# Nutri-Score labelling in the out-of-home food sector: evidence on effectiveness from a large, randomised control trial in UK restaurant settings

**DOI:** 10.64898/2026.07.29.26359233

**Authors:** Amy Finlay, Andrew Jones, Rebecca Evans, Zoé Colombet, James Garbutt, Rosanna May Maletta, Martin O’Flaherty, Zoi Toumpakari, Nick Townsend, Eric Robinson

**Affiliations:** Department of Psychology, University of Liverpool, UK; School of Psychology, Liverpool John Moores University, UK; Department of Public Health, Policy and Systems, University of Liverpool, UK; Department for Health, University of Bath, UK; School for Policy Studies, University of Bristol, UK; School of Psychology, University of Greater Manchester, UK

## Abstract

**Background:** Over half of UK adults eat food prepared out-of-home (OOH) weekly and the poor nutritional profile of OOH food contributes to ill health. Nutri-Score is a form of interpretative labelling which assigns food products a value of A (healthiest) to E (least healthy) based on nutritional quality, and inclusion on menus could be a public health policy option to reduce obesity. This is the first real-world Randomised Control Trial to test the effectiveness of Nutri-Score alongside calorie labelling on food menus in UK OOH settings.

**Methods:** Adult participants (N=672), majority White (80%) female (63%), with a mean age of 47(±18) years, were recruited from the local community to visit OOH food businesses to order, consume, and pay for meals. Data collection days were randomised to be control (calorie labelling) or Nutri-Score (calorie labelling with Nutri-Score). Linear mixed models assessed impacts of labelling condition on perceived effectiveness of menu labelling and nutritional quality of food orders.

**Findings:** Scores for perceived effectiveness of labelling were significantly greater in the Nutri-Score condition compared to control (B = 0.21, p=0.010; 95% CI 0.05, 0.37) and food orders were significantly better nutritional quality, indicated by lower Nutrient Profiling Model scores in the Nutri-Score vs. control condition (B = -0.89, p=0.003; 95% CI -1.48, -0.31).

**Interpretation:** Compared to providing calorie labelling alone, inclusion of Nutri-Score could be an effective policy approach to improve nutritional quality of diet and reduce diet-related disease.

**Funding:** This research was funded by the Economic and Social Research Council (ESRC;Reference ES/W007932/1).

**Research in context:** *Evidence before this study:* PubMed was searched for all research into Nutri-Score food labelling, published between June 2016 (earliest available result), and February 24, 2026, using the search terms: (((((Nutri-Score)) OR (Nutriscore)) OR (Nutri-Score label*)) OR (Nutriscore label*)) with no language restrictions. The search returned n=354 articles deemed relevant according to title and abstract content. Nutri-Score labelling has been adopted as a front-of-pack food labelling policy in several European countries. Systematic reviews and meta-analyses have explored the impact of Nutri-Score labelling, primarily on food packaging, through both real world and experimental evidence. Findings suggest that Nutri-Score is effective in helping consumers to identify healthier foods, and rank foods in order of healthiness. Additionally, Nutri-Score labelling is associated with improved nutritional quality of food selection, by facilitating the selection of healthier food choices and the avoidance of less-healthy food choices. However, most studies to date have been conducted in experimental settings, and only n=2 studies have tested Nutri-Score in out-of-home (OOH) settings. The two studies conducted in the OOH food sector were conducted in France and differed in design (e.g., non-randomized). Findings have suggested that Nutri-Score is associated with food choices of greater nutritional quality, although effects may not be consistent across socioeconomic status.

*Added value of this study:* No research has examined the effectiveness of Nutri-Score on food choices with a UK population or the impact of Nutri-Score on nutrient consumption (as opposed to purchase or selection). Inclusion of interpretive labelling (i.e., Nutri-Score) for OOH food menus is a potential avenue for national government policy, however research to assess effectiveness and guide implementation of such policy is limited. In the present real-world randomised controlled trial, participants visited an OOH food business in Liverpool city centre and selected a meal from a control menu (calorie labelling only) or a Nutri-Score intervention menu (calorie labelling with Nutri-Score). Primary outcomes of interest were perceived message effectiveness and nutritional quality of meal ordered.

*Implications of all the available evidence:* The evidence suggests that Nutri-Score labelling combined with calorie labelling on food menus is perceived to be effective by consumers and may prompt healthier food choices than calorie labelling alone. The inclusion of interpretative nutrition information, such as Nutri-Score, should be considered for OOH settings as a public health policy.

## Background

In the UK, it is estimated that 60% of adults eat food prepared outside of the home (OOH) weekly^(1)^ and similar trends of regular OOH food consumption are observed internationally^(2)^. Such foods are typically higher in fat, salt, and sugar and lower in fibre and fruit and vegetable content than foods prepared at home^(3)^, and are associated with increased daily energy intake^(4)^. Because greater consumption of OOH food is associated with poorer overall dietary quality, the growing influence of the OOH food sector on population-level diet is a significant public health challenge.

Food labelling strategies can be used to inform consumers of the nutritional content of foods and promote healthier food choices. Labelling interventions can also prompt food reformulation by the food industry^(5)^. In England, during 2022, it became mandatory for large OOH food businesses (with more than 250 employees) to provide calorie content information for menu items. This policy does not appear to have resulted in OOH consumers reducing energy consumed during OOH visits^(6)^, but may have promoted a small amount of product reformulation^(7)^. As part of a formal policy post-implementation review, the English government will decide whether to continue to mandate calorie labelling alone or introduce a revised menu labelling system for OOH food.

Nutri-Score labelling was initially implemented as a front-of-pack food labelling policy in France, and has since been introduced as a voluntary front-of-pack labelling scheme in several other European countries^(8)^. Nutri-Score is a form of interpretative labelling which assigns food products an A (healthiest) to E (least healthy) category. Category allocation is based on overall nutritional quality, assessed via the content of energy, saturated fat, salt, sugar, fibre, protein and fruit, vegetable and nut content.

Systematic reviews and meta-analyses have explored the impact of Nutri-Score labelling through real-world and experimental evidence, primarily on food packaging. Findings suggest that Nutri-Score is effective as a front of pack label compared to other labelling strategies (e.g. multiple traffic light labelling, health star ratings) for helping consumers to identify healthier foods. Nutri-Score also performs well in aiding ranking of foods by nutritional quality for individuals of all levels of socioeconomic position (SEP)^(9)^. A network meta-analysis of majority (95%) lab-based research found that Nutri-Score labelling compared to no labelling was associated with a reduced odds of selecting less healthy products (Odds Ratio: 0.66; 95% CI 0.53, 0.82), and increased overall nutritional quality of food selection by 7.9% (95% CI 1%, 14%)^(10)^. However, research on the potential application of Nutri-Score to OOH settings is limited and this could be an important avenue for policy development.

Only two studies have examined Nutri-Score labelling in OOH settings^(11, 12)^. One study implemented Nutri-Score labelling in a workplace cafeteria in France as part of a non-randomized pre-post observational design without a control group^(12)^. Inclusion of Nutri-Score on menus was associated with an improvement in the overall nutritional quality of food ordered, as measured by Nutrient Profiling Model (NPM) scores, and a reduced selection of total energy, sugar, and saturated fat^(12)^.

However, this study did not collect information on consumption and was not able to examine differential effects of Nutri-Score by participant demographic characteristics. A second study introduced Nutri-Score labelling on the menu of a French fast food chain serving sandwiches over three separate experiments^(11)^, one of which was conducted in real-world settings. Nutri-Score (vs. no nutrition information) was associated with healthier food choices in both hypothetical and real-world settings by facilitating consumer’s identification of healthier options. However, improvements to nutritional quality of food choices were only consistently shown for middle-class individuals living in metropolitan areas^(11)^. If replicated in other countries, this social patterning of consumer response to Nutri-Score may be less desirable because greater or equivalent impacts on diet are needed in groups of lower SEP to reduce well observed inequalities in diet-related ill health^(13)^.

Interpretive labelling such as Nutri-Score for OOH food menus is likely of interest to policymakers because of the limited impact existing policy options (e.g., calorie labelling alone) may have. Consistent with this, the French government are currently considering the use of Nutri-Score in OOH settings^(11)^. Inclusion of Nutri-Score alongside calorie content may be acceptable to the public as Nutri-Score provides a more global assessment of nutritional quality ^(14)^. Nutri-Score is also easily understood by consumers^(9)^ whereas there is limited public understanding of calorie labels^(15)^. However, research to guide the implementation of Nutri-Score OOH is limited and it is not clear whether inclusion of Nutri-Score alongside existing mandated energy content (as in the US, England and other countries) would lead to healthier food choices.

In the present study we used a randomised controlled trial design in which participants visited OOH businesses, ordering from menus with vs. without the inclusion of Nutri-Score. Primary outcomes of interest were perceived message effectiveness (PME) of menu information in motivating healthier food behaviours and nutritional quality of meal ordered. Hypotheses were as follows:

**H1:** Participants in the Nutri-Score (intervention; calorie labelling with Nutri-Score) condition will score the menu as having higher PME compared to participants in the control (calorie labelling only) condition.

**H2:** Participants in the Nutri-Score (intervention) condition will order a meal with greater nutritional quality (indicated by lower UK NPM scores) compared to participants in the control condition.

## Methods

### Study design and participants

This study was a between-subjects randomised controlled trial conducted in two OOH businesses in Liverpool city centre (England, UK). One business was a ‘typical’ English café serving foods such as sandwiches, jacket potatoes, fish and chips. The second business was a vegan restaurant, serving breakfast and lunch foods including pancakes, sandwiches, burgers and salad bowls. The study was pre-registered on the Open Science Framework (https://osf.io/tm7vw) and Clinical Trials (NCT number: NCT06923241). The study received ethical approval from the University of Liverpool Ethics committee (reference: 4612).

Participants were recruited through an existing database of local volunteers at the University of Liverpool and social media advertising targeted to the local area between the 27^th^ March 2025 and the 31^st^ July 2025. Participants were required to complete a screening questionnaire, and were eligible to take part in the study if they were over the age of 18, a fluent English speaker, not currently pregnant or breastfeeding, regularly ate food prepared OOH (at least once a month), and were able to visit a restaurant in Liverpool city centre for lunch on their own or with up to four known acquaintances. All participants who completed the study were reimbursed £20 for travel and time commitments associated with participation.

Participants were asked to ensure their acquaintances also met the inclusion criteria. Follow-up data revealed that a small number of participants (6% of total) did not meet the criteria ‘regularly ate food prepared OOH’ (at least once a month) but were retained in primary analysis. This is a deviation from the pre-registered analysis plan, however removal of these participants did not change the statistical significance of any primary analysis results.

Recruitment was stratified to obtain a sample representative of the UK population in terms of gender, age, and highest level of education. Figure 1 is a consort flow diagram depicting study participation.

**Figure 1:**
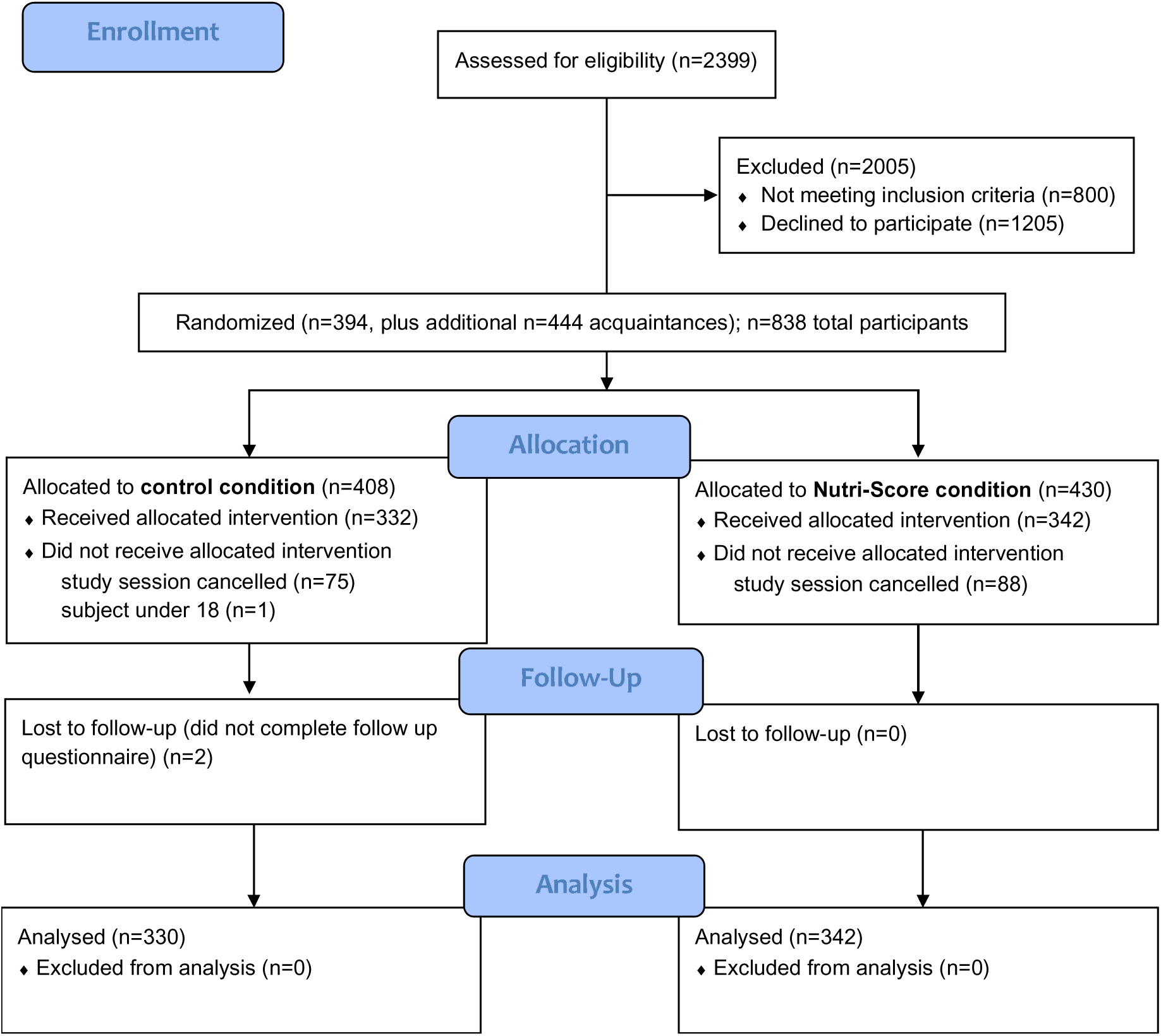
Consort flow diagram of study participation.

### Study randomisation

Study data collection days were randomised to be either the control condition (calorie labelling only) or Nutri-Score condition (calorie labelling with Nutri-Score). Calorie labelling information was included in the control menus as large OOH food businesses in England are required by law to provide this information on menus and we aimed to examine whether inclusion of Nutri-Score alongside calorie labelling may be an effective policy approach. Randomisation occurred at the level of study day (Monday-Friday) to prevent intervention contamination (e.g., participants seeing the menu of a different condition) and conducted by researchers using the RAND function in Excel. Participants were blind to study condition. For full information on allocation ratio, see online supplementary materials (A). The study was advertised as ‘consumer behaviour in different restaurants’ to disguise the true aims of the research, and participants were informed of the full study aims upon debriefing.

### Study Setting

We selected OOH businesses with menu items that were anticipated to have a wide range of calorie content and nutritional quality, covering Nutri-Score values A-E. Researchers worked with the outlet staff to gather the necessary information to calculate energy and nutrient content and Nutri-Score values for all menu items.

Existing menus used by businesses were adapted for the study and approved by business owners as being feasible to implement. See online supplementary materials (B) for all study menus and details of gathering nutritional information. Reference information for calorie intake (‘Adults need on average 2000 kcal per day’) and Nutri-Score (short description of how Nutri-Score is calculated) were provided on menus according to menu condition.

### Calculating Nutri-Score and energy content

Nutri-Score was calculated using the 2024 approach provided by the Nutritional Epidemiology Research Team, Paris Nord University^(16)^. This accounts for a food’s energy (kj), saturated fat (g), sugar (g), salt (g), fibre (g), protein (g), and fruit, vegetable and nut content (%) per 100g/100ml, as well as the presence of non-nutritive sweetener (Y/N) in drinks.

Nutri-Score does not currently account for portion sizes, and this is a limitation to its application to the OOH food sector. We therefore used lower and upper thresholds for energy content and daily limits for nutrients based on UK nutrition guidance(17–19) to adjust Nutri-Score gradings. This resulted in a small number of menu items being upgraded to a healthier grading (e.g., from C to B) and downgraded to a less healthy grading (from C to D). For further detail of adjustments to Nutri-Score, see online supplementary materials (C).

### Procedure

Participants attended a lunch session at one of two outlets scheduled between 12pm and 3pm Monday to Friday. Participants were welcomed, given a verbal summary of the study information (which had been emailed to them in advance), and asked to provide verbal consent before being given study menus. Research staff were trained by the individual businesses to act as servers (e.g., take orders, deliver food to tables, clear plates).

Prior to food being served to tables and after clearing tables, images of food and drink were covertly taken by research staff in the outlet kitchen in order to estimate consumption. Once plates were cleared, participants were given a QR code to complete the follow-up questionnaire on their own device, or on a research team tablet. Follow-up questionnaires included questions on demographics, participant characteristics, menu labelling and study aim guessing. Once completed, participants were asked to pay for their meal and told they would need to complete a dietary recall questionnaire the following morning.

The following morning participants were emailed with a link to complete a 24-hour dietary recall using a validated tool; Intake24^(20)^. Participants reported all food and drink eaten after the meal, until midnight on the same day to explore whether nutrient intake after the OOH visit differed between conditions. Participants were then fully debriefed.

### Primary outcome measures

#### Perceived Message Effectiveness

To measure perceived effectiveness of Nutri-Score labelling by consumers, an adapted version of the University of North Carolina Perceived Message Effectiveness (PME) scale was used^(21)^. The 3-item scale uses a 5-point Likert scale and mean scores are calculated. Higher scores indicate higher PME. PME has been frequently used to assess perceived consumer benefit of food labelling interventions and is predictive of behaviour(^21, 22^).

#### Nutritional quality of meal

Nutritional quality of ordered meal was calculated using the UK Nutrient Profiling Model (NPM 2004/05^(23)^) scoring system which Nutri-Score is based on. The UK NPM has been used in prior research to evaluate dietary and meal healthiness and is based on the UK dietary guidelines so is directly applicable to the study sample. Scores are calculated for each product by deducting the total score for positive nutrients from the total score for negative nutrients. Lower scores (minimum -15) indicate greater nutritional quality, while higher scores (maximum +40) indicate poorer nutritional quality^(24)^.

It was anticipated that participants would order multiple items as part of a meal (e.g. a main dish, a side and a drink) and therefore to obtain a single nutritional quality score for the full meal, mean UK NPM scores were calculated, weighted according to the energy content of the item, as in previous research^(12)^.

### Secondary outcome measures

#### Nutri-Score label of selected menu items

Participants were coded as having ordered any of the healthiest menu items labelled A or B (Y/N) and/or the least healthy menu items labelled D or E (Y/N).

#### Energy and Nutrient content of food ordered/consumed

The energy content of all menu items ordered by participants were recorded. To estimate consumption, images of food were taken before serving to participants, and once participants had finished eating. Using these images, and available information on the proportion made up by each dish component, researchers estimated the percent of each dish consumed by participants. See online supplementary materials (D) for an example of images taken and calculation. Two researchers individually estimated the percentage eaten and a mean percentage was taken as the final estimate.

Participants also reported whether any foods were shared with other participants and the proportion shared, and whether anything was added to their meal (e.g. salt, pepper, condiments). This information was used to estimate the energy and nutrients consumed over the meal by each participant.

### Other Measures

#### Later energy and nutrient intake

Using Intake24, participants reported all food eaten from the end of the study session until the end of the day (midnight that night). Total energy and nutrient (salt, fat, saturated fat, sugar, protein, fibre) intake were used to test for any evidence of later compensation after the study intervention.

#### Demographics

Participants reported their age, gender, height and weight (to calculate BMI), ethnicity, and highest level of education achieved.

#### Health Food Choice Motives

The health subscale of the Food Choice Motives Questionnaire^(25)^ was used to measure the extent that individuals prioritise health when making food choices.

#### Label awareness/understanding

Participants were asked if they saw any nutritional information on the menu they ordered from. If they selected ‘Yes’ they were asked to specify from a list what nutritional information they saw: Nutri-Score, Traffic light labelling, Nutrient warning labels, Keyhole healthy choices, Calorie information, or none of the above options. Images of labels accompanied each of the label options.

All participants were then shown an image of the outlet menu they ordered from with Nutri-Score and calorie labels. They were asked if the Nutri-Score labels influenced (Nutri-Score condition) or would have influenced (control condition) their food choices, and if the answer was ‘Yes’ asked to specify how their choice was influenced or would have been influenced. Finally, participants were asked if they would support a government policy that required the implementation of Nutri-Score on food menus.

### Analysis

A pre-registered analysis strategy was followed (https://osf.io/tm7vw), and all analyses were conducted in R (version 4.5.2). For R packages used, see Supplementary Materials (E). In all models, outlet visited (plant-based/traditional), gender (female/male & other), SEP (low education = no degree/high education = degree level and above), and health food choice motives (continuous) were included as fixed effects. The intervention was considered a fixed effect and a random slope was specified at the cluster level. Analyses were clustered by study day (Monday-Friday) and table (i.e., participant group ranging from 1-5 members), so these variables were included as random effects. However, in preliminary model fitting, study day contributed no variance to one model and produced a perfect negative correlation in another model. As a result, day was removed as a random effect and condition removed as a random slope in final models which had negligible effect on model outcomes. A comparison of results from primary models with day included vs excluded as a random effect are reported in Supplementary Materials (F).

Results for primary analyses were considered significant at p<0.025 (adjusted due to interim analysis, see pre-registration) and secondary analyses at p<0.010.

#### Primary analyses

Two linear mixed regression models were used to assess the impact of labelling condition on the primary outcomes of PME (continuous) and nutritional quality of ordered meal as measured by mean weighted NPM scores (continuous).

#### Secondary analyses

Two logistic mixed regression models were used to assess whether labelling condition was associated with the odds of selecting a healthier (A/B labelled) item, or a less healthy (D/E labelled) item. Further linear mixed models assessed whether labelling condition was associated with total energy and individual nutrient content (salt, sugar, fat, saturated fat, protein, fibre, fruit vegetable and nut content) of food consumed.

#### Exploratory analyses

To explore any evidence of later compensatory eating behaviour, linear mixed models explored differences in later energy and nutrient intake between the two conditions.

In a second step of the two primary linear models, participant characteristics (age, gender, education and health food choice motives) were included as interaction terms to test for moderation of intervention condition by participant characteristics. Continuous predictor variables were mean centred. Outlet was also tested as a moderator. This final moderation analysis was not pre-registered in error and has not impacted any other planned analyses.

#### Sensitivity analyses

Primary analyses were repeated with participants who guessed the aim of the study (n=47) removed. We also assessed whether our primary findings remained the same when nutritional quality of the ordered meal was replaced with nutritional quality of the consumed meal.

#### Sample size and Interim analyses

In pre-registered power calculations prior to data collection, we estimated we would require at least N=450 participants. Due to intervention effect size uncertainty, we conducted pre-planned interim analyses at n=294, before using effect size information and evidential certainty (Bayes Factors) at interim to determine that collecting further data would be informative and what the likely sample size required to improve evidential certainty would be^(26)^. Based on this approach, our final target sample size was N=700. See online supplementary materials (G) for full information.

## Results

A full study flow diagram is depicted in Figure 1. A total of N=672 participants were included in the final analysis. The majority of participants were White (80%), female (63%), frequent consumers of OOH food (94% once a week or more) with a mean age of 47 years. There was a slightly greater proportion of high SEP individuals compared to low SEP (58% vs. 42%), and the average BMI of all participants was 26.2. Participant characteristics overall and by condition are reported in Table 1.

**Table 1:** Participant characteristics in the overall sample, and by condition.

|  | Overall (n=672) | Control (n=330) | Nutri-Score (n=342) |
| --- | --- | --- | --- |
|  | N (%) | N (%) | N (%) |
| **Gender** |  |  |  |
| Female | 424 (63%) | 204 (62%) | 220 (64%) |
| Male/other | 248 (37%) | 126 (38%) | 122 (36%) |
| **SEP\*** |  |  |  |
| Low education | 280 (42%) | 152 (46%) | 128 (37%) |
| High education | 392 (58%) | 178 (54%) | 214 (63%) |
| **Ethnicity** |  |  |  |
| White | 541 (80%) | 270 (82%) | 271 (79%) |
| Asian | 86 (13%) | 39 (12%) | 47 (14%) |
| Mixed | 20 (3%) | 11 (3%) | 9 (3%) |
| Black | 15 (2%) | 6 (2%) | 9 (3%) |
| Other | 10 (2%) | 4 (1%) | 6 (1%) |
| **OOH food consumption frequency** |  |  |  |
| Less than once per month | 40 (6%) | 18 (6%) | 22 (6%) |
| 1-3 times per month | 408 (61%) | 199 (60 %) | 209 (61%) |
| 1-2 times per week | 188 (28%) | 96 (29%) | 92 (27%) |
| 3 times per week or more | 36 (5%) | 17 (5%) | 19 (6%) |
|  | **Mean (standard deviation)** | **Mean (standard deviation)** | **Mean (standard deviation)** |
| **Age** | 46.61 (18.01) | 48.22 (18.50) | 45.06 (17.42) |
| **BMI\*\*** | 26.16 (5.11) | 26.42 (5.38) | 25.90 (4.82) |
| **Health Food Choice Motives** | 2.77 (0.66) | 2.79 (0.68) | 2.75 (0.65) |

\*SEP = socioeconomic position; Low education = no degree, high education = degree-level and above
\*\*BMI = Body Mass Index

Details of participant food orders are reported in full in Table 2. The mean weighted NPM scores for ordered meals was 0.39 and the total energy ordered by participants was on average 885kcal. Further detail of nutritional quality of meals ordered and consumed split by outlet is shown in online supplementary materials (H).

**Table 2:**
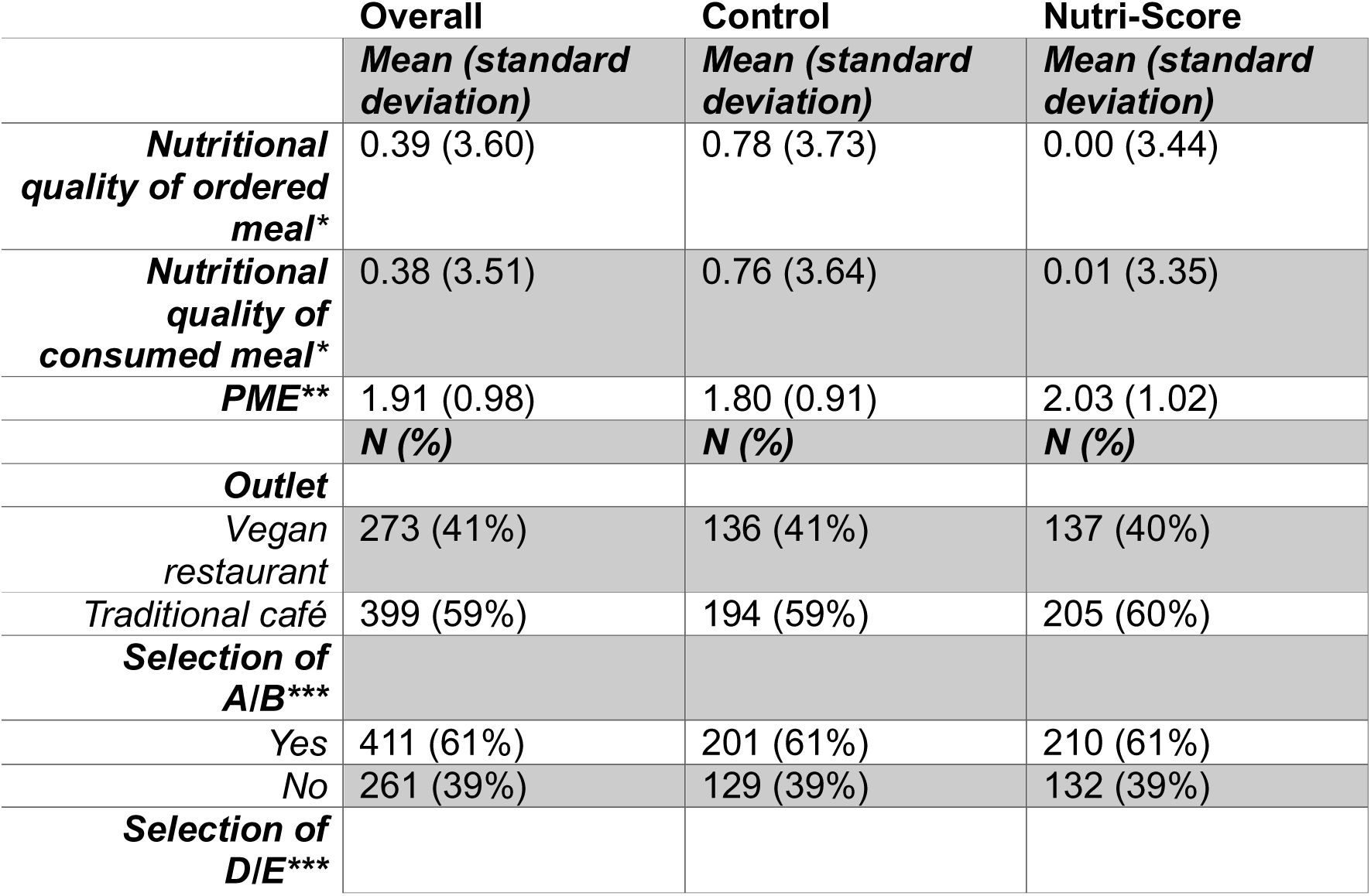
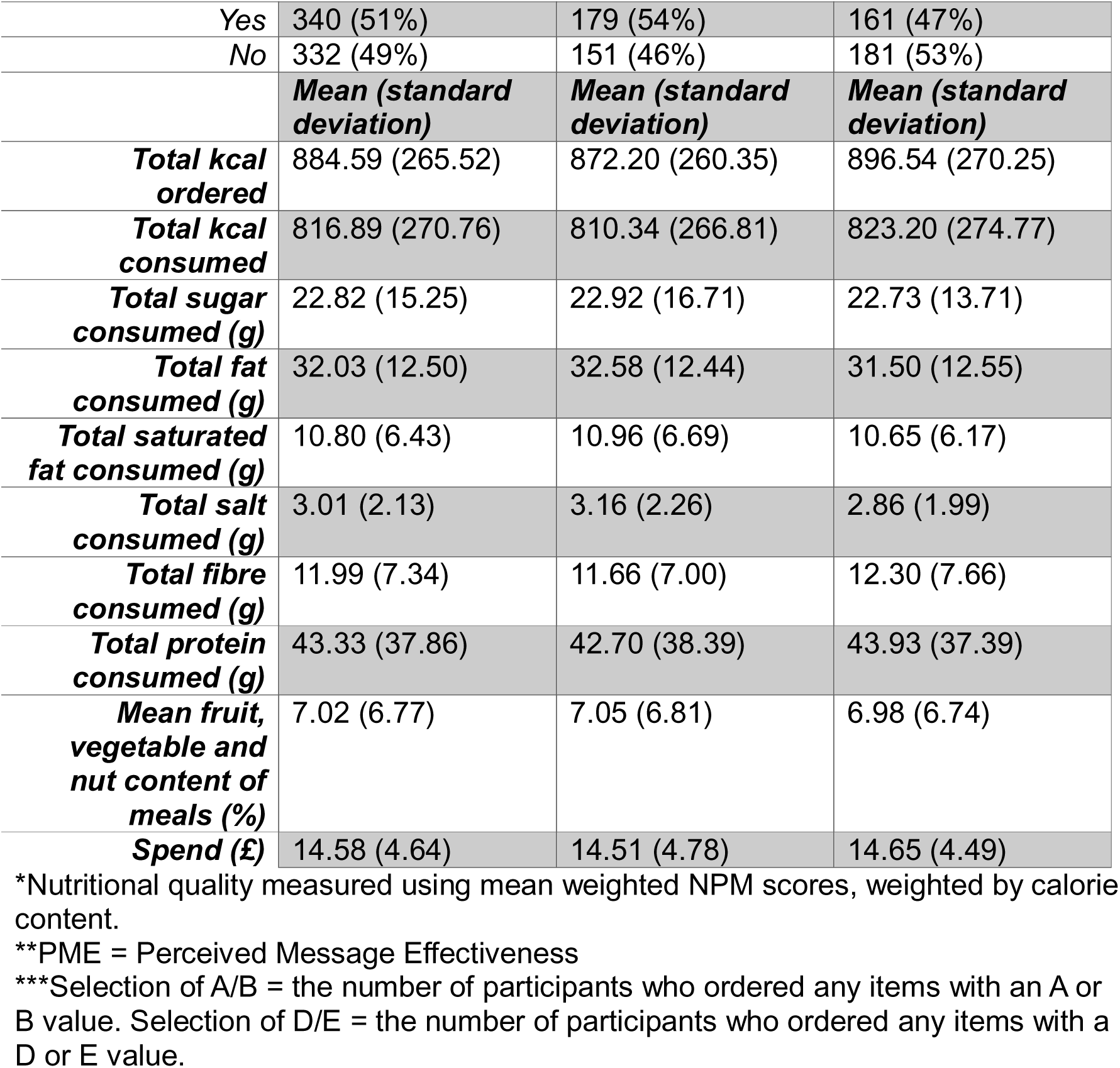
Food order details overall and by condition.

### Primary outcomes

The Nutri-Score condition was associated with significantly greater PME than the control condition (B = 0.21, p=0.010; 95% CI 0.05, 0.37, see Figure 4), with Bayes Factors calculated using a simplified t-test indicating moderate evidence for an effect of intervention (BF^10^ = 7.7). The Nutri-Score labelling condition also led to significantly lower weighted NPM scores (B = -0.89, p=0.003; 95% CI -1.48, -0.31) indicating greater nutritional quality of selected meals (See Figure 5), with Bayes Factors calculated using a simplified t-test indicating moderate evidence for an effect of intervention (BF^10^ = 4.1).

**Figure 4:**
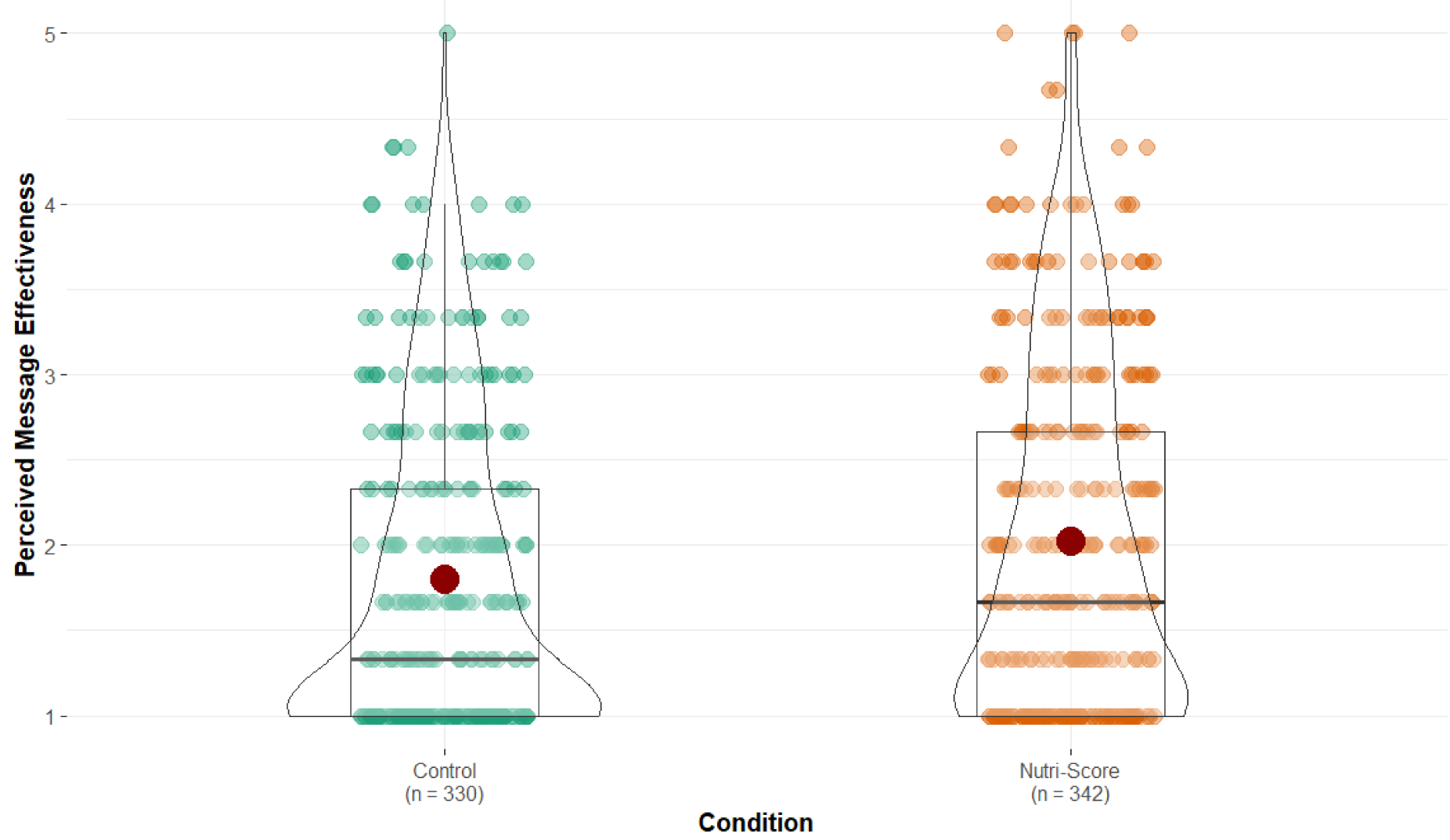
Violin plot showing between group comparisons of PME. *Boxes show the median and interquartile range of observations, while the mean is shown by red dots. The shape of the violin plot represents the density of data (i.e. a wider section of the plot = more data points).

**Figure 5:**
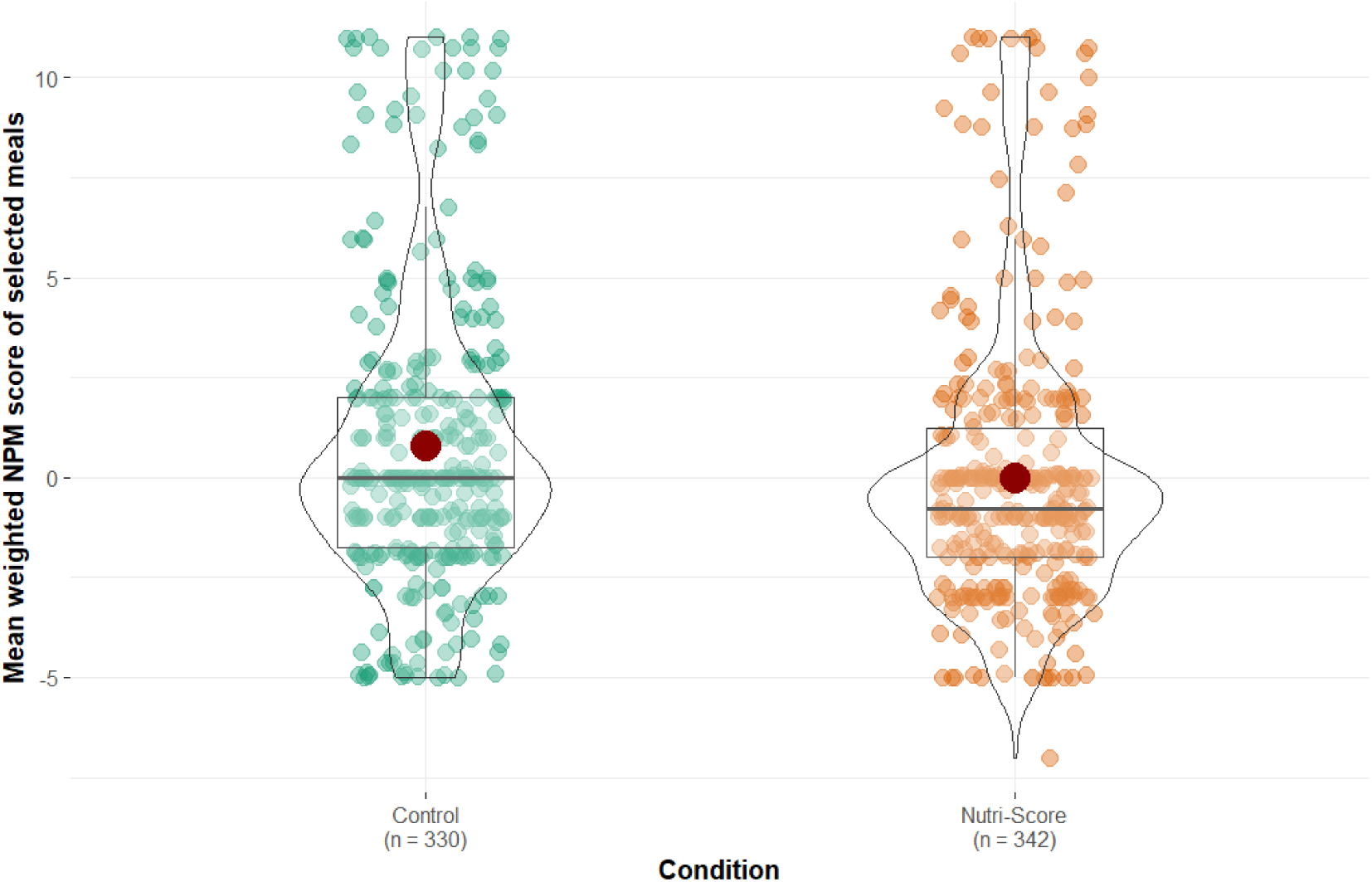
Violin plot showing between group comparisons of meal nutritional quality, measured by mean NPM scores weighted by energy content. *\*Boxes show the median and interquartile range of observations, while the mean is shown by red dots. The shape of the violin plot represents the density of data (i.e. a wider section of the plot = more data points). NPM = weighted Nutrient Profiling Model scores*.

### Secondary outcomes

A logistic regression model showed that there was no significant difference between the two labelling conditions in the odds of selecting at least one healthier item (labelled A/B: (aOR = 1.07, p=0.733; 95% CI 0.73, 1.55), with a 0.5 percentage point difference between conditions. Similarly, there was no significant difference in the odds of selecting at least one less healthy item (labelled D/E) between the two conditions (aOR = 0.69, p=0.060; 95% CI 0.47, 1.02), with selection of D/E menu items 7 percentage points lower in the Nutri-Score condition vs. Control.

#### Energy and Nutrient intake

Mixed linear models, with the same adjustments as primary analyses, showed no observed associations between the two labelling conditions for energy or any individual nutrient intake. See Table 2 and online supplementary materials (I) for analyses in full.

#### Exploratory analyses

##### Later intake of energy and nutrients

Later intake of energy and nutrients collected through dietary recall assessment tool Intake24 overall and by condition is shown in online supplementary materials (J). No significant associations between study condition and later energy or nutrient intake were observed.

##### Moderation analyses

There was no evidence of interactions of age, gender, education, health food choice motives or outlet with condition on the two primary outcome variables (see online supplementary materials (K)).

#### Sensitivity analyses

Primary findings were the same when participants who guessed study aims (n=47) were removed. We also assessed whether the primary findings remained the same when nutritional quality of the ordered meal was replaced with nutritional quality of the consumed meal, and results remained the same (see online supplementary materials (L)).

#### Questionnaire responses

The majority of participants (69%) stated that if the UK government introduced a policy requiring Nutri-Score on food menus they would support or strongly support it.

Most participants in the Nutri-Score condition correctly reported the presence of Nutri-Score on the menu they ordered from (68%). A minority of participants in the Nutri-Score condition (38%) reported that the Nutri-Score information influenced their choice. See online supplementary materials (M) for questionnaire response results in full.

#### Additional post-hoc analyses

As we observed a significant improvement in overall nutritional quality of food ordered and consumed in the Nutri-Score condition, but no significant increase/decrease in likelihood of ordering at least one A/B (healthiest) or D/E (least healthy) menu item (planned analyses), we further descriptively explored ordering patterns to better understand differences between conditions. As displayed in online supplementary materials (N), there were small directional tendencies for participants in the Nutri-Score condition (vs. Control) to only order food items that were the healthiest on menus (13% relative increase) and not the least healthy (12% relative decrease). In further post-hoc analyses, we repeated our two primary linear models split by outlet type and there was a tendency for Nutri-Score to have a directionally (but not statistically significantly) larger impact on outcomes in the traditional café compared to vegan restaurant outlet (see online supplementary materials (H)).

## Discussion

The inclusion of Nutri-Score labelling on food menus in OOH food outlets was associated with greater PME and healthier food orders compared to control condition menus which included calorie information only. While overall nutritional quality of meals was higher, there were no statistically significant effects of condition on the likelihood of selecting any of the healthiest (A/B Nutri-Score category) or least healthy (D/E Nutri-Score category) menu items. Additionally, observed changes to the overall nutritional quality of meals was not driven primarily by an increase or decrease in a specific nutrient or energy content of purchased meals.

The observed increase in perceived effectiveness of provided nutrition information on menus attributable to Nutri-Score is consistent with the use of Nutri-Score resulting in participants becoming more concerned about the health effects of consuming less healthy menu items. Perceived effectiveness measures have frequently been used to test the effectiveness of health messaging relating to nutrition labelling policies^(27)^. Longitudinal research suggests that greater PME is predictive of long-term health behaviour change such as weight loss^(28)^. The PME results of the present study are consistent with consumers finding Nutri-Score labelling in OOH settings helpful^(9)^ and this could be predictive of longer-term behavioural change.

The inclusion of Nutri-Score labelling resulted in the meals ordered by participants being of better overall nutritional quality compared to control, which is consistent with the PME outcome. Mean weighted NPM scores were ∼0.82 lower on average in the Nutri-Score labelling condition. This size of effect may be meaningful at population level or over repeated eating out occasions. Previous research found that for scores of overall dietary quality (as opposed to a single meal), an increase in NPM score of one point was associated with a 16% higher obesity risk in men^(29)^. In the present study, exploratory analyses suggested inclusion of Nutri-Score labelling resulted in no change to later dietary intake after the OOH outlet visit. Therefore, effects of Nutri-Score on OOH food consumption could have positive effects on overall dietary quality, and if sustained, long-term health. Future research will now be needed to examine the longer-term effects of Nutri-Score labelling in the OOH food sector.

Despite the overall improvement in the nutritional quality of meals ordered in the Nutri-Score condition, there were no statistically significant differences between conditions in the likelihood of selecting at least one menu item classed as healthier (A/B), or less-healthy (D/E). Given there were not large differences in consumption of energy or any one nutrient, improved overall nutritional quality of orders was likely driven through a combination of small but cumulative changes across nutrients.

Previous research has proposed that Nutri-Score labelling works by discouraging less healthy choices^(11)^, and while a trend toward a reduction in the likelihood of any less healthy menu item choices (7%) was observed in the Nutri-Score condition, our study was not powered for these secondary analyses. Observed differences with prior research may be due to the differences in menu options between the two studies. The most comparable study on Nutri-Score used a menu from a single sandwich shop chain in France. In comparison, the present research examined behaviour across two different UK outlet types with a wide selection of food types, across all Nutri-Score classifications. It is possible that greater variation in cuisine types, cultural differences and/or study designs may help explain differences between studies.

Previous research has observed a reduction in energy selection in response to Nutri-Score labelling^(12)^, however, no such effect was observed in the present study. It is possible this is because in the current study, Nutri-Score was implemented alongside calorie labelling. While previous research has implemented Nutri-Score on food menus as a standalone label(^11, 12^), the aim of the present research was to test whether Nutri-Score had an effect greater than calorie labelling alone which currently exists as an OOH food policy in England.

A greater proportion of participants noticed nutritional information in the Nutri-Score condition vs the control condition. This is consistent with previous research demonstrating that colour-coded labels can be effective in prompting healthier choices as they are eye-catching and aid understanding of nutritional information^(10)^. Most participants (69%) stated that they would support the implementation of Nutri-Score labelling on food menus, suggesting that Nutri-Score would be well accepted by the public were it to be implemented alongside calorie labelling. However, the implementation of a calorie labelling policy in England resulted in discussion around the potential negative impacts on individuals with eating disorders^(30)^ and there is little understanding currently of how Nutri-Score may affect individuals with eating disorders. It is possible that Nutri-Score could reduce the likelihood of harm as it considers the overall nutritional quality of a food product rather than focusing on calories. Unlike a recent study in France^(11)^, we found no evidence that effects of Nutri-Score differed by individual-level participant socioeconomic status (education level).

### Strengths and limitations

This is the first randomized controlled trial to examine whether inclusion of Nutri-Score labelling alongside calorie information in OOH settings impacts real-world behaviour and the first trial to examine Nutri-Score in the OOH food sector outside of France. However, as this study was conducted in two outlets in Liverpool, UK, the results may not be generalisable to other outlet types or wider socio-demographic groups. Consistent with this, we found some directional evidence that the impact Nutri-Score had on PME and nutritional quality of orders was numerically larger in the traditional café vs. vegan restaurant outlet, which may reflect differences in menus and/or the types of participants who chose to eat at each outlet. Participants were recruited through social media advertising and a database of local research volunteers who wanted to engage in research. Therefore, our sample may not be representative of the typical population. We also cannot generalise findings to other ethnicities as participants were predominantly White British.

Previous research identified that Nutri-Score influenced food choices only in middle-class individuals living in metropolitan areas^(11)^. As the present study was conducted in a city centre, our results are likely not generalisable to individuals living in more rural areas and more formal testing of this with UK consumers may now be warranted. Limitations of the present study also include a lack of data on longer-term impacts of Nutri-Score on consumer behaviour. Whilst we examined consumer behaviour in real-world settings, participants were aware they were participating in a study. Further observational research examining effects on consumer behaviour after implementation of Nutri-Score in out of home settings would therefore be valuable. A final limitation is that while later compensation was measured through dietary recall, we did not explore compensation that may have occurred prior to the study session in anticipation of a meal.

## Conclusions

The inclusion of Nutri-Score combined with calorie labelling (vs. calorie labelling alone) on food menus was associated with greater perceived message effectiveness and healthier food choices in OOH settings. Compared to the US and England’s requirement of provision of calorie information alone in OOH settings, the inclusion of Nutri-Score labelling could be an effective policy approach to improving population level diet.

## Data Availability

All data produced in the present study are available online at https://osf.io/tm7vw/

## Acknowledgments

We would like to acknowledge the help of the owners and employees of the businesses used for this research: The Waterhouse Café and The Vibe in Liverpool city centre. We would also like to acknowledge Emily Rizk, Isla Finlay and Francesca Quigley for their help with data collection.

## References

1. Mariani E, Chacko A, Stewart I, et al. How eating out contributes to our diets. UK: Nesta; 2024.

2. Lachat C, Nago E, Verstraeten R, et al. Eating out of home and its association with dietary intake: a systematic review of the evidence. Obesity reviews. 2012;13(4):329–46.

3. Gesteiro E, García-Carro A, Aparicio-Ugarriza R, et al. Eating out of Home: Influence on Nutrition, Health, and Policies: A Scoping Review. Nutrients. 2022;14(6):1265.

4. Garbutt J, Townsend N, Johnson L, et al. The contribution of the out-of-home food (OOHF) sector to the national diet: a cross-sectional survey with repeated 24-hour recalls of adults in England (2023-2024). medRxiv. 2025:2025.06.30.25330369.

5. Packer J, Michalopoulou S, Cruz J, et al. The Impact of Non-Fiscal Mandatory and Voluntary Policies and Interventions on the Reformulation of Food and Beverage Products: A Systematic Review. Nutrients. 2024;16(20):3484.

6. Polden M, Jones A, Essman M, et al. Evaluating the association between the introduction of mandatory calorie labelling and energy consumed using observational data from the out-of-home food sector in England. Nature Human Behaviour. 2025;9(2):277–86.

7. Essman M, Burgoine T, Huang Y, et al. Changes in energy content of menu items at out-of-home food outlets in England after calorie labelling policy implementation: a pre–post analysis (2021–2022). BMJ Public Health. 2025;3(2).

8. Dréano-Trécant L, Egnell M, Hercberg S, et al. Performance of the Front-of-Pack Nutrition Label Nutri-Score to Discriminate the Nutritional Quality of Foods Products: A Comparative Study across 8 European Countries. Nutrients. 2020;12(5):1303.

9. Shrestha A, Cullerton K, White KM, et al. Impact of front-of-pack nutrition labelling in consumer understanding and use across socio-economic status: A systematic review. Appetite. 2023;187:106587.

10. Song J, Brown MK, Tan M, et al. Impact of color-coded and warning nutrition labelling schemes: A systematic review and network meta-analysis. PLOS Medicine. 2021;18(10):e1003765.

11. Chandon P, Indaburu A. When and how simplified nutrition labels improve fast-food choices. Journal of the Academy of Marketing Science. 2026.

12. Julia C, Arnault N, Agaësse C, et al. Impact of the Front-of-Pack Label Nutri-Score on the Nutritional Quality of Food Choices in a Quasi-Experimental Trial in Catering. Nutrients. 2021;13(12):4530.

13. Løvhaug AL, Granheim SI, Djojosoeparto SK, et al. The potential of food environment policies to reduce socioeconomic inequalities in diets and to improve healthy diets among lower socioeconomic groups: an umbrella review. BMC Public Health. 2022;22(1):433.

14. Polden M, Robinson E, Jones A. Assessing public perception and awareness of UK mandatory calorie labeling in the out-of-home sector: Using Twitter and Google trends data. Obesity Science & Practice. 2023;9(5):459–67.

15. Krukowski RA, Harvey-Berino J, Kolodinsky J, et al. Consumers May Not Use or Understand Calorie Labeling in Restaurants. Journal of the American Dietetic Association. 2006;106(6):917–20.

16. Recherche en Epidémiologie Nutritionelle. Spreadsheet to calculate the updated version of the Nutri-Score n.d. [Available from: https://nutriscore.blog/2022/12/25/spreadsheet-to-calculate-the-updated-version-of-the-nutri-score/.

17. Better Health. Calorie counting UK: NHS; n.d. [Available from: https://www.nhs.uk/better-health/lose-weight/calorie-counting/.

18. Public Health England. Behind the headlines: calorie guidelines remain unchanged. UK; 2017.

19. Public Health England. Calorie reduction: Technical report: guidelines for industry, 2017 baseline calorie levels and the next steps. UK: GOV.UK; 2020.

20. Bradley J, Simpson E, Poliakov I, et al. Comparison of INTAKE24 (an Online 24-h Dietary Recall Tool) with Interviewer-Led 24-h Recall in 11–24 Year-Old. Nutrients. 2016;8(6):358.

21. Noar SM, Barker J, Bell T, et al. Does Perceived Message Effectiveness Predict the Actual Effectiveness of Tobacco Education Messages? A Systematic Review and Meta-Analysis. Health Communication. 2020;35(2):148–57.

22. Grummon AH, Hall MG, Taillie LS, et al. How should sugar-sweetened beverage health warnings be designed? A randomized experiment. Preventive Medicine. 2019;121:158–66.

23. Department of Health. Nutrient Profiling Technical Guidance.; 2011.

24. Bowes Byatt L, Bunting T. How should we define healthy food in policy? The case for the nutrient profile model: Nesta; n.d. [Available from: https://www.nesta.org.uk/toolkit/how-should-we-define-healthy-food-in-policy/.

25. Steptoe A, Pollard TM, Wardle J. Development of a measure of the motives underlying the selection of food: the food choice questionnaire. Appetite. 1995;25(3):267–84.

26. Kairalla JA, Zahigian R, Wu SS. Interim Analysis in Clinical Trials. In: Piantadosi S, Meinert CL, editors. Principles and Practice of Clinical Trials. Cham: Springer International Publishing; 2022. p. 1083–102.

27. Taillie LS, Hall MG, Popkin BM, et al. Experimental Studies of Front-of-Package Nutrient Warning Labels on Sugar-Sweetened Beverages and Ultra-Processed Foods: A Scoping Review. Nutrients. 2020;12(2):569.

28. Lehto T, Oinas-Kukkonen H. Explaining and predicting perceived effectiveness and use continuance intention of a behaviour change support system for weight loss. Behaviour & Information Technology. 2015;34(2):176–89.

29. Julia C, Ducrot P, Lassale C, et al. Prospective associations between a dietary index based on the British Food Standard Agency nutrient profiling system and 13-year weight gain in the SU.VI.MAX cohort. Preventive Medicine. 2015;81:189–94.

30. Frances T, O’Neill K, Newman K. ‘An extra fight I didn’t ask for’: A qualitative survey exploring the impact of calories on menus for people with experience of eating disorders. British Journal of Health Psychology. 2024;29(1):20–36.

