## Supplementary Material for "Nutri-Score labelling in the out-of-home food sector: evidence on effectiveness from a large, randomised control trial in UK restaurant settings"

**Supplementary Materials**

**A: Randomisation & allocation ratio**

Randomisation was initially allocated (by day) at a 1:1 ratio for intervention vs. control using RAND function in Microsoft Excel. However, after 11 weeks of data collection, the proportion of participants in the control vs intervention groups was imbalanced. We calculated the total number of participants required in each condition to achieve a balanced (approximately 1:1) sample for remaining study days and for the final 6.5 weeks of data collection randomisation at a ratio of 3:1 (Control:Intervention).

**B: Creating study menus**

For the vegan restaurant, the online data management software ‘Nutritics’(1) was used to calculate the energy and nutrient content of menu items by the research team. For the traditional café, the outlet staff routinely used software ‘ProcureWizard’(2) to record ingredients, recipes and allergens. The inputted data was checked by research staff, and the same platforms were used to calculate energy and nutrient content of menu items. All menus used in the study are shown in Figures 1-6.

**Figure 1: Traditional café menu for the Nutri-Score condition (calorie labelling with Nutri-Score)**

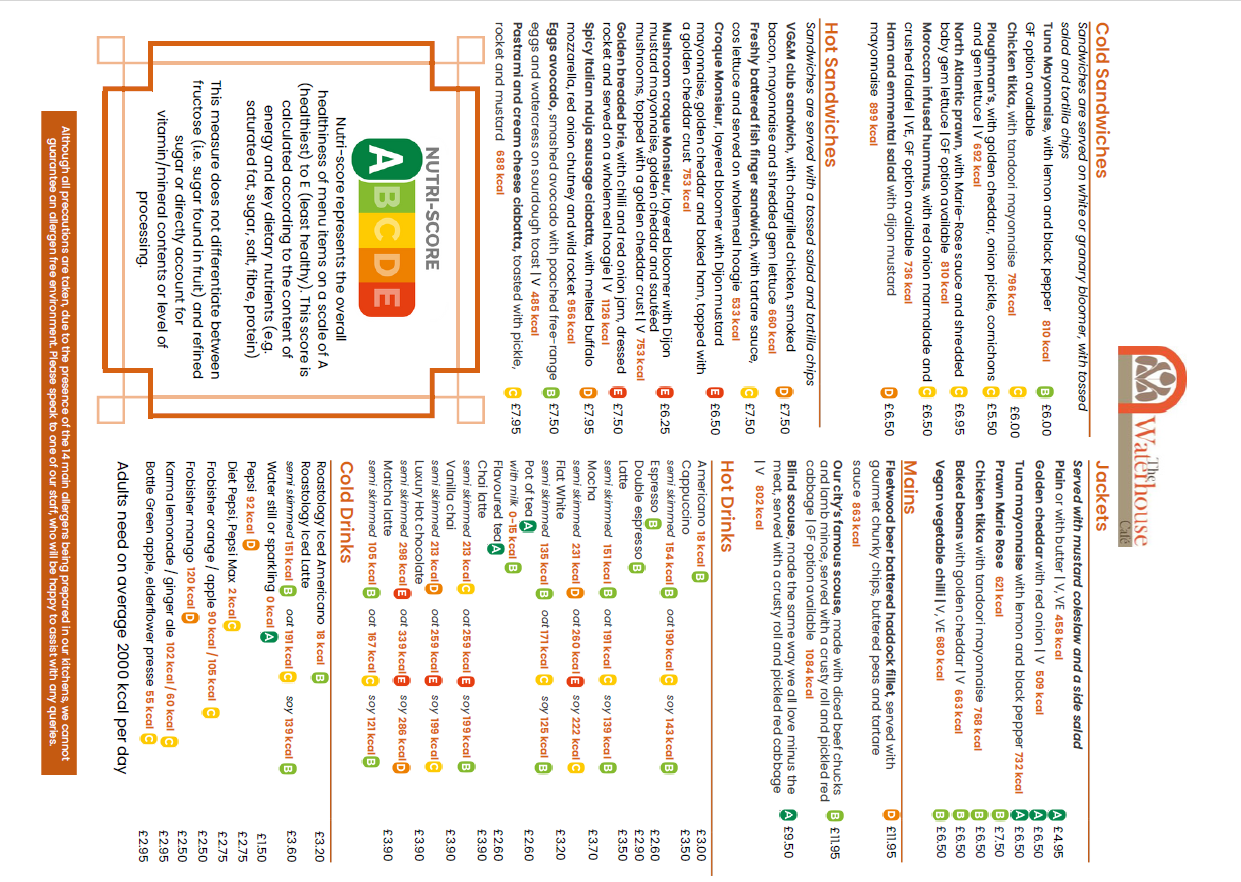

**Figure 2: Traditional café menu for control condition (calorie labelling only)**

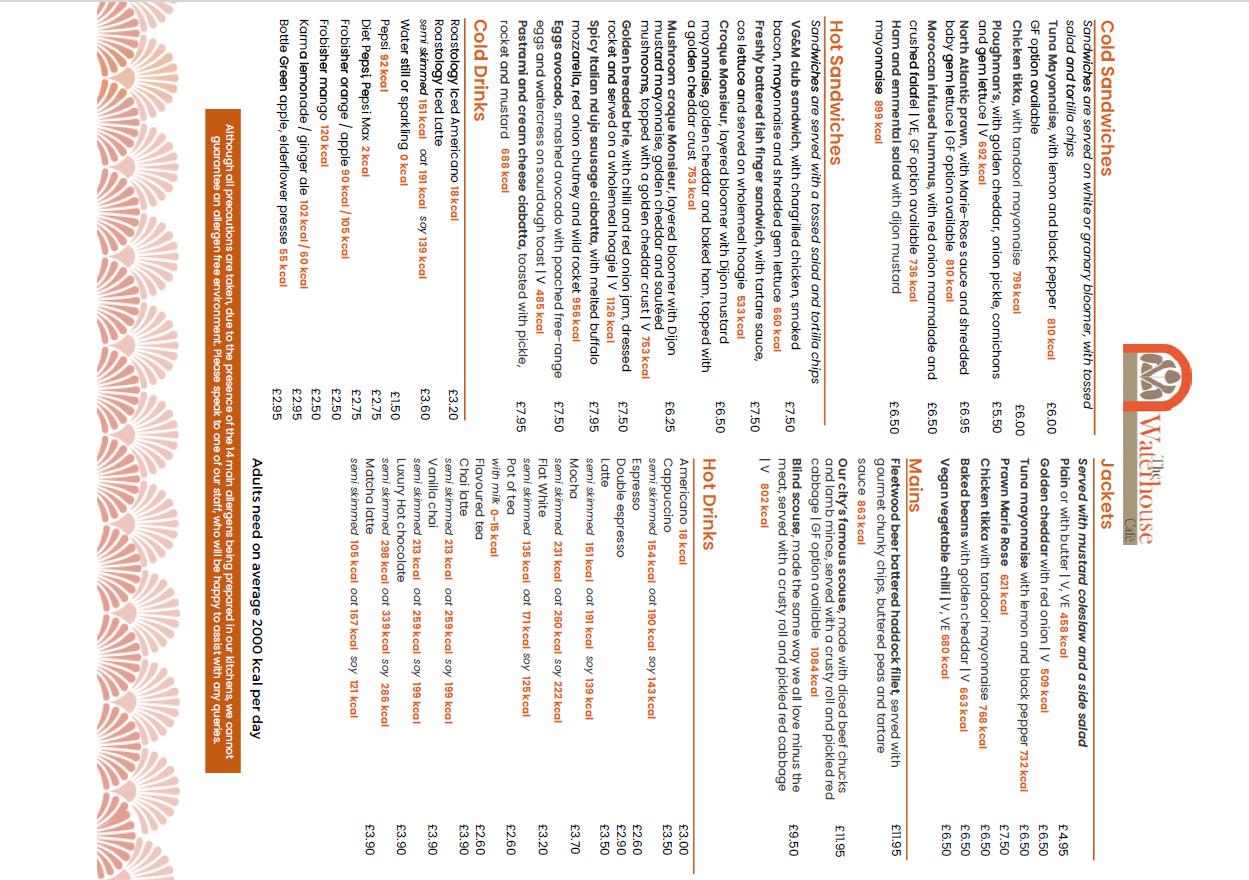

**Figure 3: Vegan restaurant menu for the Nutri-Score condition (calorie labelling with Nutri-Score) (front)**

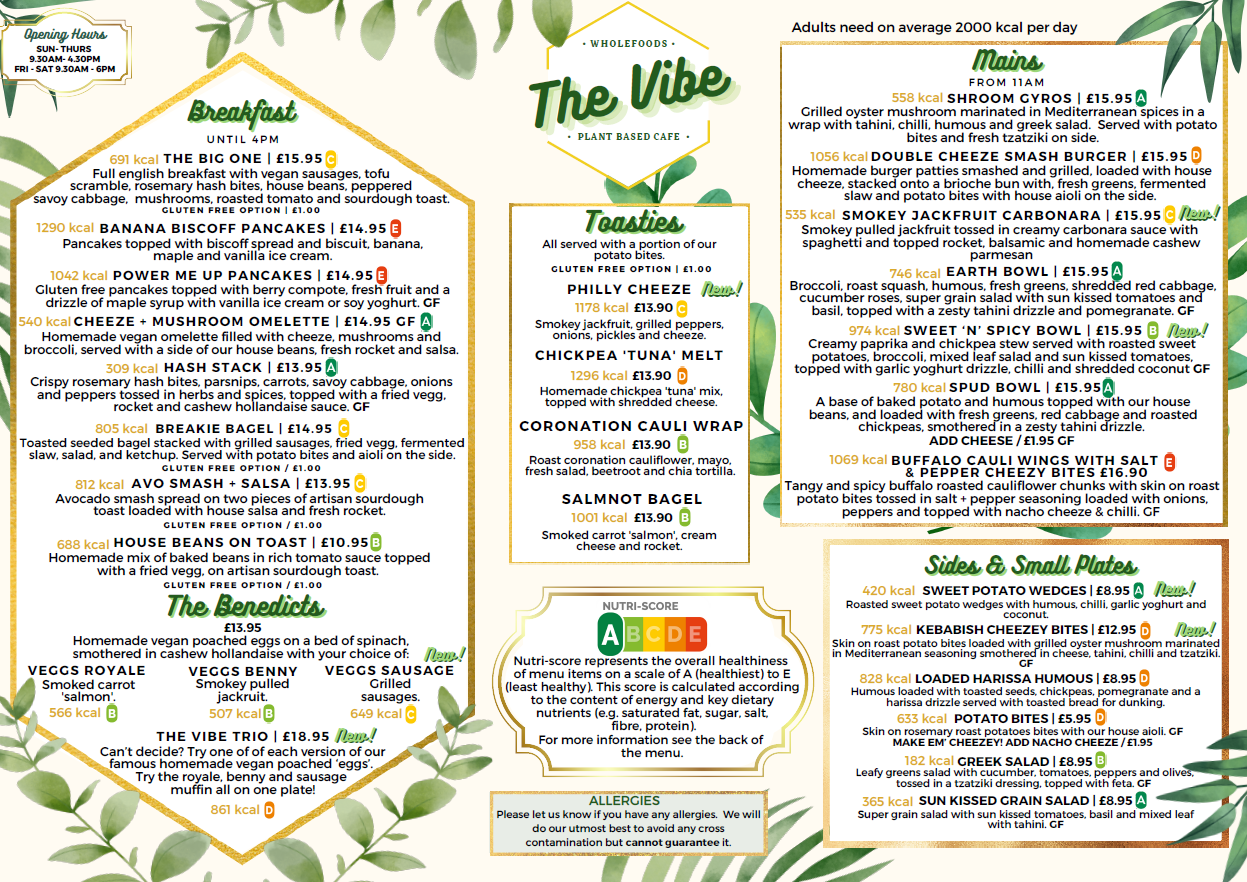

**Figure 4: Vegan restaurant menu for the Nutri-Score condition (calorie labelling with Nutri-Score) (back)**

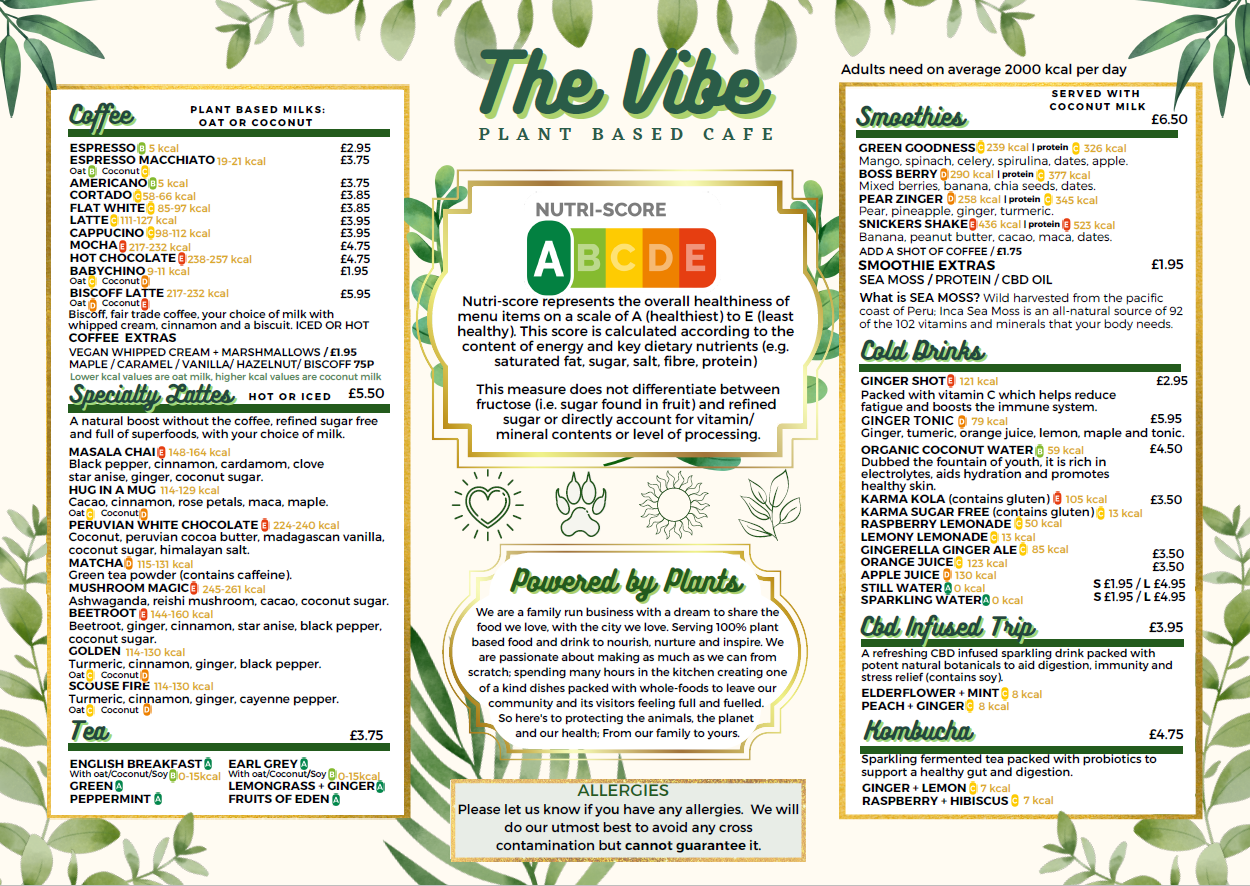

**Figure 5: Vegan restaurant menu for control condition (calorie labelling only)**

**(front)**

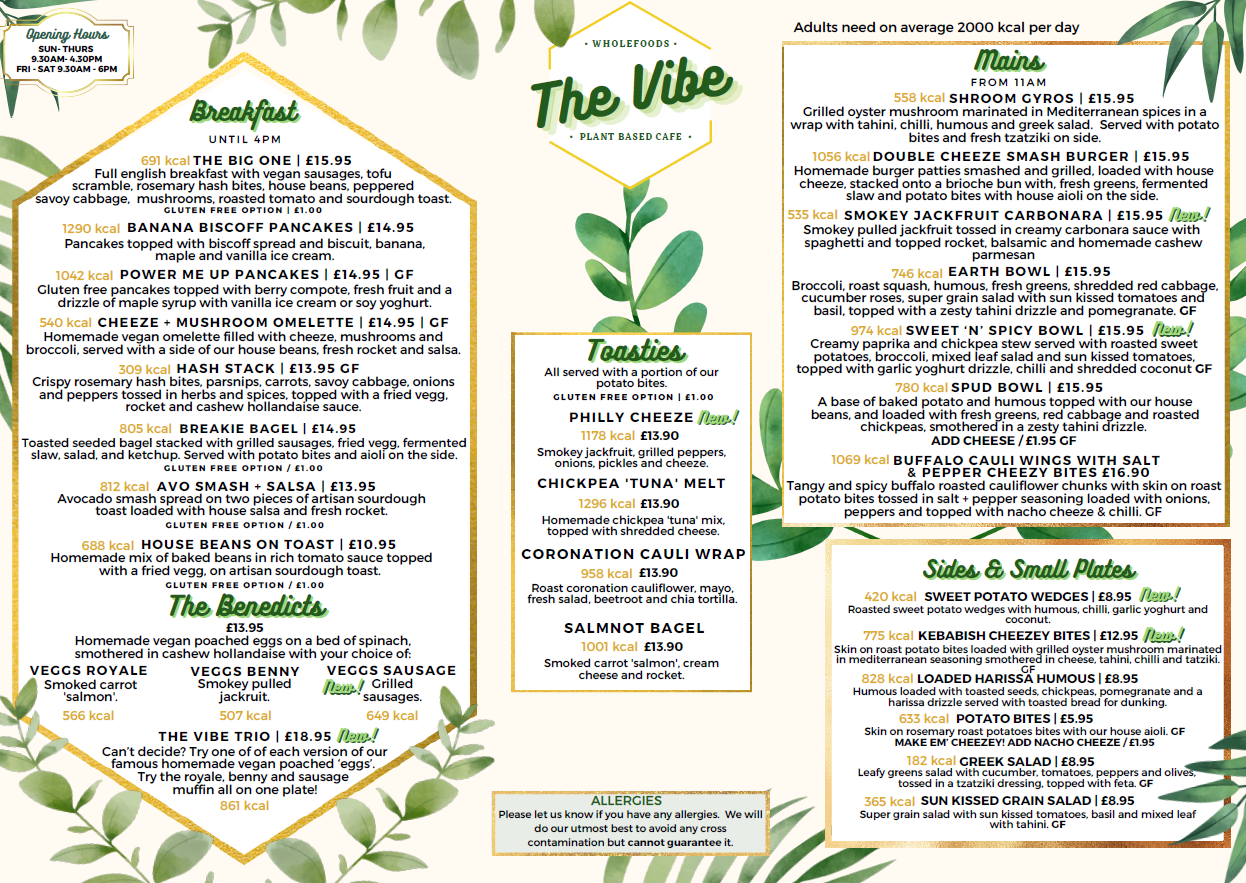

**Figure 6: Vegan restaurant menu for control condition (calorie labelling only) (back)**

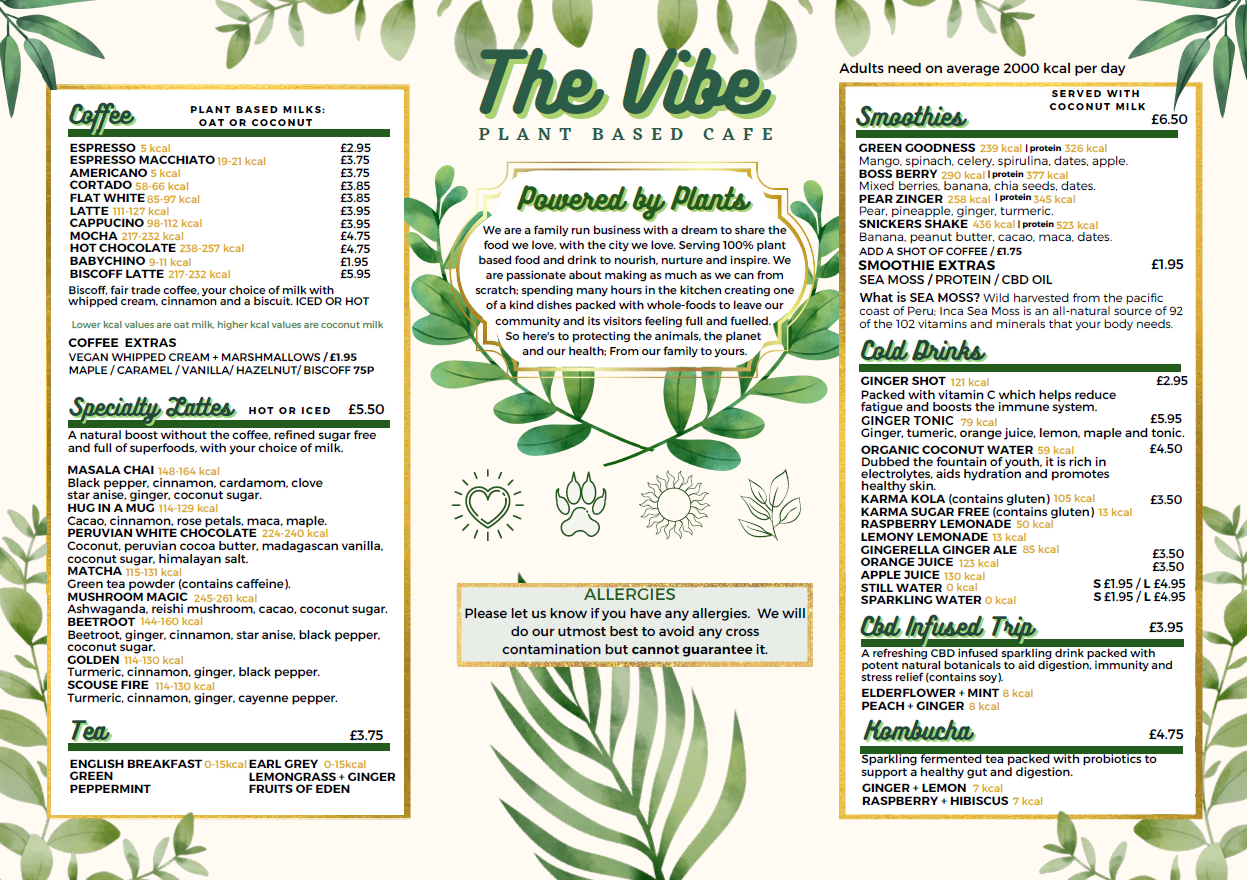

**C: Adjustments to Nutri-Score**

To account for portion size and total amount of energy for menu items, we operationalised lower and higher energy content thresholds based on UK dietary and out-of-home food sector guidelines:

- Starters and sides 280kcal (lower threshold) and 600kcal (higher threshold)
- Mains 600kcal (lower threshold) 860kcal (higher threshold)

280kcal and below (lower threshold) was deemed a suitable size for a starter or side dish to accompany a main meal, based on NHS guidance for snacks(3). Six hundred kcal and below was selected as the lower threshold based on recommended energy content for main meals(4). We determined starters and sides having as much or more energy than a recommended main (600kcal) would constitute an excessive amount (higher threshold for starters/sides). We classed above 860 kcal (higher threshold for mains) as excessive for main meals, as it would exceed the target energy content of a main meal outlined by the UK calorie reduction technical report (5).

If the energy content of menu items was below the lower threshold value, items were upgraded by a single Nutri-Score category (e.g. from C to B). If the energy content of menu items was above the higher cut off value, items were downgraded a single Nutri-Score category (e.g. from C to D). Additionally, if any items contained ≥100% of the average guideline daily allowance of energy (2000kcal), saturated fat (20g), salt (6g), or sugar (30g) as recommended by UK government, the menu item was classed as the least healthy NS category (E).

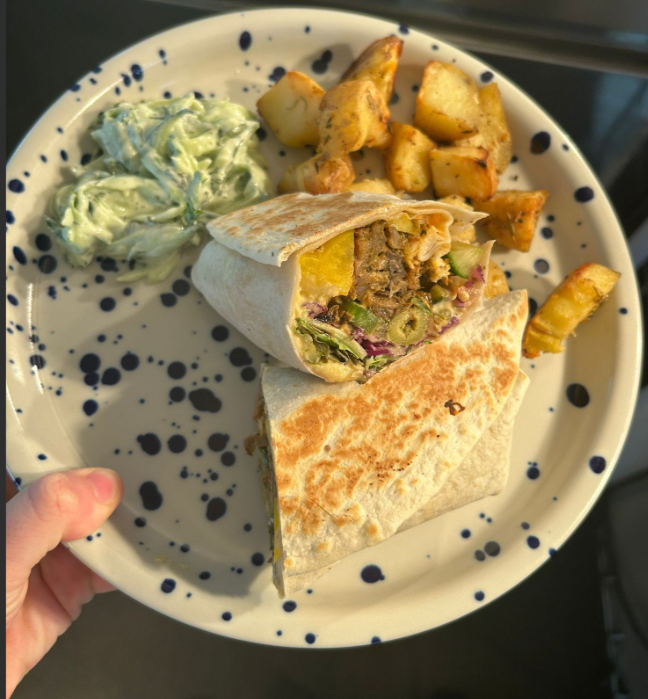

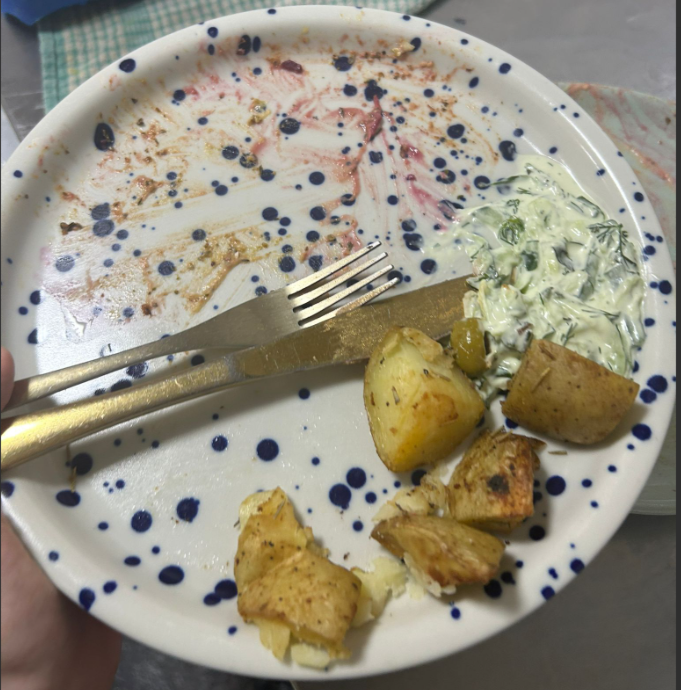
**D: Example images used to calculate consumption**

| **Shroom gyros** | **Grams** | **% of dish weight** | **% eaten** |
| --- | --- | --- | --- |
| Tzatziki | 15 | 3 | 1 |
| Pomegranate flesh and pips | 10 | 2 | 2 |
| Greek salad | 75 | 16 | 16 |
| Cabbage red raw | 30 | 7 | 7 |
| Hummus | 30 | 7 | 7 |
| Mushroom gyros mix | 120 | 26 | 26 |
| Gyros wrap | 56 | 12 | 12 |
| Hash bites | 120 | 26 | 14 |
|  |  | **SUM:** | **85%** |

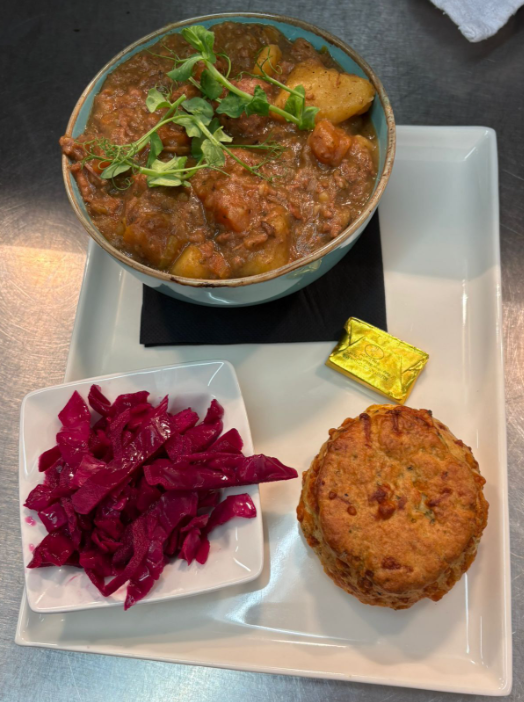

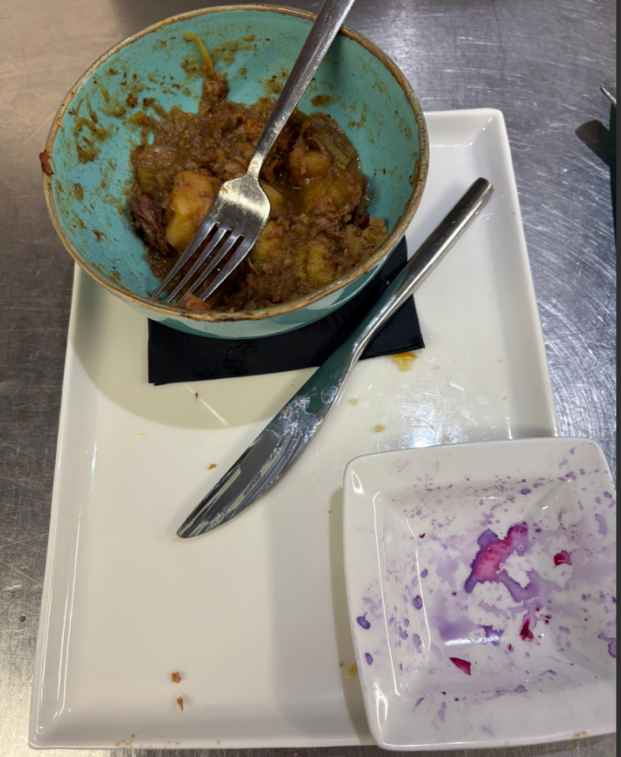

| **Scouse** | **Grams** | **% of dish weight** | **% eaten** |
| --- | --- | --- | --- |
| Cheese scone/butter | 104 | 12 | 12 |
| Red cabbage | 50 | 6 | 6 |
| Scouse | 687 | 82 | 60 |
|  |  | **SUM:** | **78%** |

**E: R packages used for analysis**

The following R packages were used: ‘mice’(28) for multiple imputation of Body Mass Index (BMI) (n=60), Perceived Message Effectiveness (PME) (n=1), and later intake (n=66); ‘ggstatsplot’ (29) for the creation of Figures; ‘lme4’ (30) for executing multi-level models.

**F: Supplementary Table 1: Results from linear mixed models with study day included as a random effect compared to final models with day removed**

| Model outcome | Day included as a random effect (planned analysis) | Day removed as a random effect (final analysis) |
| --- | --- | --- |
| PME* | 0.21, p=0.010; 95% CI 0.05, 0.37 | 0.21, p=0.010; 95% CI 0.05, 0.37 |
| Weighted NPM** scores | -0.82, p=0.007; 95% CI -1.41, -0.23 | -0.89, p=0.003; 95% CI -1.48, -0.31 |
| Likelihood of any A_B | 1.10, p=0.758; 95% CI 0.61, 1.97 | 1.07, p=0.733; 95% CI 0.73, 1.55 |
| Likelihood of any D_E | 0.71, p=0.181; 95% CI 0.43, 1.17 | 0.69, p=0.060; 95% CI 0.47, 1.02 |

*PME = Perceived Message Effectiveness **NPM = Nutrient Profiling Model

**G: Power calculation and interim analyses**

Prior to data colllection we conducted a simulation to estimate sample size required for two primary outcomes: Perceived Message Effectiveness (PME) and mean weighted Nutrient Profiling Model (NPM) scores. Our simulation was based on previous research and knowledge of the outlets confirmed for data collection.

We simulated running the study with 20 participants a day for 20 days (400 total data points). For PME, we estimated from previous research that we may have an intercept PME value of 1.85 (SD: 1.08), with an intervention effect of 1 (i.e., the intervention group will have PME values 1 unit greater than the control group on average, based on previous research(52)), and minimal difference in PME scores between the two outlets (0.2). We also assumed ICC of approximately 0.200, and a standard deviation of the random intercept of eating group in a mixed model of 1. Across 200 simulations using a regression model clustered by day, we would have 94% power (95% CI 89.75, 96.86) to detect differences in PME between the two conditions.

For NPM scores, we calculated using the two confirmed outlet menus, that we may have an intercept NPM score of -0.27 (SD: 3.29) with an intervention effect of 0.22 (the intervention group will select meals with greater nutritional quality, shown by a 0.22 higher NPM score) based on previous research(26) and a mean difference between the two outlets of 1.4 (the difference in mean NPM scores for the two outlets). We also assumed an ICC of approximately 0.819, and a standard deviation of the random intercept of eating group in a mixed model of 1. Across 200 simulations using a regression model clustered by day we would have 88.5% power (95% CI 83.25, 92.57) to detect differences in NPM scores between the two conditions.

Due to the uncertainty of effects, and lack of directly comparable evidence, we increased the pre-data collection target sample size (n=400) by 12.5% to n=450 participants and planned to conduct interim analyses at approximately 50% of the target sample size.

When over 50% (n=294) of the required sample size had been recruited, as in our pre-registered analysis protocol, interim analyses were conducted to assess whether convincing evidence of an intervention effect was already present at interim, and if not, whether it would be justifiable to continue data collection. At interim, there was little evidence of an effect for PME (Cohen’s d = -0.19; 95% CI -0.43, 0.06) and the evidence was in favour of the null hypothesis (BF10 = 0.05). Evidence for nutritional quality was weakly in favour of the alternative hypothesis (BF10 = 1.08) and there was an intervention effect size of d=0.21; 95% CI -0.03, 0.46). Because there was no convincing evidence of either an absence or presence of an intervention effect, we used simulation modelling to determine the sample size required to detect the size of intervention effects observed at interim (d=0.2, approximately). This indicated that approximately n=640 participants would be required to detect an effect of the intervention condition. To account for any participant exclusions for analyses, we aimed to recruit a final sample size of N=700 participants.

**H: Supplementary Table 2: Nutritional quality (weighted NPM score) for meals ordered and consumed split by outlet and condition.**

|  | Overall (M(SD)) | Control (M(SD)) | Nutri-Score (M(SD)) |
| --- | --- | --- | --- |
| **Nutritional quality of ordered meal** | | | |
| Vegan restaurant | -0.80 (3.01) | -0.71 (3.21) | -0.90 (2.82) |
| Traditional café | 1.20 (3.75) | 1.83 (3.73) | 0.61 (3.68) |
| **Nutritional quality of consumed meal** | | | |
| Vegan restaurant | -0.80 (2.90) | -0.70 (3.06) | -0.89 (2.75) |
| Traditional café | 1.18 (3.66) | 1.79 (3.67) | 0.60 (3.57) |

** Nutritional quality score for vegan restaurant = 0.17, Nutritional quality score for traditional café = 0.47*

**Supplementary Table 3:** **Results of primary linear models repeated separately for each outlet.**

| Variable | PME | Mean weighted NPM |
| --- | --- | --- |
| Primary analysis (n=672) | 0.21, p=0.010; 95% CI 0.05, 0.37 | -0.89, p=0.003, 95% CI -1.48, -0.31 |
| Vegan restaurant (n=273) | 0.06, p=0.646; 95% CI -0.19, 0.31 | -0.21, p=0.607; 95% CI -1.01, 0.59 |
| Traditional café (n=399) | 0.32, p=0.003; 095% CI 0.11, 0.52 | -1.38, p=0.001; 95% CI -2.19, -0.56 |

**I: Supplementary Table 4: Results of linear models for associations between study condition and energy and nutrient intake over the study session**

| Nutrient* | Estimate |
| --- | --- |
| Energy (kcal) | 19.89, p=0.368; 95% CI -23.42, 63.20 |
| Sugar (g) | -0.49, p=0.719; 95% CI -3.19, 2.20 |
| Fat (g) | -0.89, p=0.367; 95% CI -2.82, 1.04 |
| Saturated fat (g) | -0.36, p=0.437; 95% CI -1.26, 0.54 |
| Salt (g) | -0.34, p=0.057; 95% CI -0.93, 0.01 |
| Fibre (g) | 0.73, p=0.194; 95% CI -0.38, 1.84 |
| Protein (g) | 0.67, p=0.816; 95% CI -4.96, 6.29 |
| Fruit, vegetable & nut content (mean %) | -0.10, p=0.845; 95% CI -1.16, 0.95 |

**Reference condition = control.*

**J: Supplementary Table 5: Later energy and nutrient intake (the rest of the day after the study session) overall and by condition.**

| Later intake* | Overall M(SD) | Control M(SD) | Nutri-Score M(SD) |
| --- | --- | --- | --- |
| Kcal | 781.06 (511.12) | 735.06 (459.12) | 825.44 (553.77) |
| Fat | 31.20 (26.12) | 28.19 (19.71) | 34.11 (30.83) |
| Saturated fat | 12.37 (12.57) | 11.90 (11.98) | 12.83 (13.13) |
| Sodium | 813.58 (693.70) | 802.45 (657.44) | 824.32 (727.77) |
| Sugar | 31.60 (35.20) | 28.97 (28.39) | 34.13 (40.58) |
| Fibre | 7.39 (6.27) | 7.08 (5.66) | 7.69 (6.80) |
| Protein | 35.57 (29.52) | 34.93 (27.67) | 36.18 (31.23) |
| Carbohydrate | 80.04 (60.50) | 74.89 (54.93) | 85.13 (65.13) |

** For later intake, missing data (n=67) were imputed.*

**Supplementary Table 6: Results of linear models for associations between study condition and later energy and nutrient intake**

| Nutrient* | Estimate |
| --- | --- |
| Energy (kcal) | 86.87, p=0.033; 95% CI 6.81, 166.94 |
| Sugar (g) | 4.77, p=0.098; 95% CI -0.89, 10.42 |
| Fat (g) | -0.89, p=0.367; 95% CI -2.82, 1.04 |
| Saturated fat (g) | 0.67, p=0.489; 95% CI -1.24, 2.59 |
| Sodium (g) | 19.53, p=0.720; 95% CI -87.43, 126.50 |
| Fibre (g) | 0.65, p=0.194; 95% CI -0.33, 1.64 |
| Protein (g) | 1.00, p=0.676; 95% CI -3.68, 5.67 |
| Carbohydrate (g) | 10.16, p=0.043; 95% CI 0.31, 20.00 |

**Reference condition = control*

**K: Supplementary Table 7: Exploration of interactions of age, gender, education, health food choice motives or outlet with condition on the two primary outcome variables**

| Interaction terms* | PME | Mean weighted NPM |
| --- | --- | --- |
| Age | -0.01, p=0.214; 95% CI -0.01, 0.00 | 0.01, p=0.441; 95% CI -0.02, 0.04 |
| Gender | 0.14, p=0.344; 95% CI -0.15, 0.42 | 0.39, p=0.466; 95% CI -0.66, 1.43 |
| Education | 0.04, p=0.802; 95% CI -0.25, 0.33 | -0.41, p=0.453; 95% CI -1.48, 0.66 |
| Health food choice motives | 0.06, p=0.574; 95% CI -0.15, 0.27 | -0.54, p=0.177; 95% CI -1.32, 0.24 |
| Outlet | -0.25, p=0.135; 95% CI -0.57, 0.08 | 1.13, p=0.061; 95% CI -0.05, 2.31 |

*reports interaction with condition in separate models

**L: Sensitivity analyses**

**Supplementary Table 8:** **Results of primary analyses models with and without participants who guessed the aim of the study**

| Variable | PME | Mean weighted NPM |
| --- | --- | --- |
| Primary analysis (n=672) | 0.21, p=0.010; 95% CI 0.05, 0.37 | -0.89, p=0.003; 95% CI -1.48, -0.31 |
| Sensitivity analysis(n=624) | 0.20, p=0.015; 95% CI 0.04, 0.37 | -0.92, p=0.002; 95% CI -1.50, -0.33 |

**Supplementary Table 9 - Replication of primary linear regression model with scores for nutritional quality of food consumed (as opposed to ordered) as the outcome variable**

| Variable | Mean weighted NPM |
| --- | --- |
| Primary analysis (Ordered) (n=672) | -0.89, p=0.003; 95% CI -1.48, -0.31 |
| Sensitivity analysis (consumed) (n=672) | -0.89, p=0.003; 95% CI -1.48, -0.31 |

**M:** **Questionnaire responses:**

Of participants in the control condition, 53% stated that if they were shown a menu with Nutri-Score, it would have influenced their decision. Of these, 23% stated they would have avoided a less healthy item and 59% stated they would have selected a healthier item. Of participants in the Nutri-Score (intervention) condition, 38% stated that the Nutri-Score labelling influenced their decision. Of these, 26% stated they avoided a less healthy item and 57% stated they would have selected a healthier item. Compared to the control condition, a greater proportion of participants in the Nutri-Score condition reported seeing any nutritional labelling (77% vs 44%). See Supplementary Table 6.

**Supplementary Table 10: The proportion of participants who noticed nutritional labelling and correctly identified the nutritional labelling they were exposed to.**

|  | Control* (n=330) N(%) | Nutri-Score** (n=342) N(%) |
| --- | --- | --- |
| Did you see any nutritional labelling? | Yes = 146 (44%)  No = 184 (56%) | Yes = 264 (77%)  No = 78 (23%) |
| Correctly identified | 130 (89%) | 77 (29%) |
| Partially correct | 5 (3%) | 182 (69%) |
| Incorrectly identified | 11 (8%) | 5 (2%) |

**For control, correct indicates that only ‘calorie labelling’ was selected. Partially correct indicates that ‘calorie labelling’ was selected alongside another label type. Incorrect indicates that ‘calorie labelling’ was not selected.
**For Nutri-Score, correct indicates that only ‘Nutri-Score’ and ‘calorie-labelling’ were selected. Partially correct indicates that either ‘Nutri-Score’ or ‘calorie labelling’ were selected alone, or with another label type. Incorrect indicates that neither ‘calorie labelling’ or ‘Nutri-Score’ were selected.*

**Supplementary Table 11: Participants’ reported support for Nutri-Score labelling as a policy in the out of home food sector**

|  | Overall (N=672) N% | Control (n=330) N% | Nutri-Score (n=342) N(%) |
| --- | --- | --- | --- |
| Strongly oppose | 13 (2%) | 8 (2%) | 5 (1%) |
| Oppose | 42 (6%) | 16 (5%) | 26 (8%) |
| Neutral | 151 (22 %) | 73 (22%) | 78 (23%) |
| Support | 275 (41%) | 132 (40%) | 143 (42%) |
| Strongly support | 190 (28%) | 100 (30%) | 90 (26%) |
| NA | 1 (<1%) | 1 (<1%) | 0 (<1%) |

**N: Additional exploratory analyses**

**Supplementary Table 12: Additional analysis of food orders, split by food and drink.**

|  | Control (N=330) | Nutri-Score (N=342) |
| --- | --- | --- |
|  | **N (%)** | **N (%)** |
| **Healthiness of drinks ordered*** | | |
| Selection of any A/B | 122 (37%) | 108 (32%) |
| Selection of any C | 127 (38%) | 141 (41%) |
| Selection of any D/E | 63 (20%) | 64 (18%) |
| **Healthiness of food ordered*** | | |
| Selection of any A/B | 127 (38%) | 154 (45%) |
| Selection of any C | 64 (19%) | 64 (19%) |
| Selection of any D/E | 143 (43%) | 130 (38%) |
| Selection of A & B **only** | 125 (38%) | 148 (43%) |
| Selection of D & E **only** | 139 (42%) | 127 (37%) |

**Participants could select multiple food dishes and drinks. N=23 participants selected two drinks, N=11 participants selected two food items.*

**References**

1.	Nutritics. Research Edition. Dublin2019.

2.	The Access Group. Access Procure Wizard Evo. n.s.

3.	Better Health. Calorie counting UK: NHS; n.d. [Available from: <https://www.nhs.uk/better-health/lose-weight/calorie-counting/>.

4.	Public Health England. Behind the headlines: calorie guidelines remain unchanged. UK; 2017.

5.	Public Health England. Calorie reduction: Technical report: guidelines for industry, 2017 baseline calorie levels and the next steps. UK: GOV.UK; 2020.
